# Retention of Female Sex Workers at one month taking oral Pre-exposure prophylaxis for HIV prevention in selected Public Health facilities in Kampala, Uganda

**DOI:** 10.1101/2022.06.28.22277029

**Authors:** James Wanyama, Christine Atuhairwe, John Bosco Alege

## Abstract

**Rationale:** Retention of Female sex workers at one month of PrEP is critical as low retention may lead to sub-optimal protection and increased HIV incidence rates in this sub-population. We determined factors associated with retention at one month among Female Sex workers on Pre-Exposure Prophylaxis for HIV prevention in selected health facilities in Kampala Capital City Authority, Uganda.

**Methods:** In this retrospective cohort study we followed 350 FSWs from September to December 2021; and 30 women underwent in-depth interviews. Retention was categorically defined as the number of FSWs who adhered to their clinic appointments got refills over the total number of FSWs expected to come for a refill. Poisson regression was used for multivariate analysis and thematic data analysis was conducted for the quantitative and qualitative data respectively.

**Results:** The median age of the FSWs was 25 years. 40.9% of the FSWs had attended primary level, 43.7% secondary level, 3.4% tertiary level education respectively, and 12% had no education. The mean duration of sex work was 3 years.On average, the FSWs had 14 sex partners in the past week. 59.4% reported lubricant use on the last sexual encounter compared to 40.6% who did not use lubricants. Factors associated with retention to PrEP at bivariate level were religion, place of work, lubricant use, use of drugs, received PrEP counselling at initiation, the color of PrEP tablet, pill size and attitude of health care workers. The Poisson regression showed that Retention to PrEP was 7% higher among sex workers that use lubricants compared to those that don’t use holding other factors constant (adjusted 0.02 [1.01, 1.14] IRR 1.07) and retention to PrEP among sex workers that think the health care workers had a good attitude was 23% higher than those that think HCW had fair attitude holding other factors constant (adjusted 0.012 [1.05, 1.44] IRR 1.23). FSWs who had no stigma had 90% higher retention than those who had stigma) (adjusted 0.04 [0.814, 0.996] IRR 0.90).No drugs at the facility affected PrEP retention by about 76% among FSWs (adjusted 0.003 [0.67, 0.922] IRR 0.76). FSWs were retained to PrEP because they felt at risk when they experienced condom breaks during intercourse or they failed to use condoms with a client. Barriers to PrEP retention included drug side effects, lack of financial resources, food insecurity, stigma, and doubts about PrEP efficacy, travel and health system factors.

**Conclusion:** A high proportion of FSWs were retained on PrEP. FSWs considered PrEP as a reliable method of protection against HIV in cases of condom breaks or if they had sex without a condom. A number of barriers to PrEP retention at both personal and systems-level need to be addressed for successful PrEP implementation.

## 1. Introduction

Globally, 37.7 million[30.2-45.1million] people were living with HIV at the end of 2020 with Key populations and their sexual partners accounting for 65% of the global HIV infections. The risk of acquiring HIV is 26 times higher for female sex workers [26], thus they are a major driver of the HIV epidemic. Approximately, 11.5%-18.6% of HIV infections in women are attributed to commercial sex work [26]. Worldwide, FSWs are 13.5 (95% CI: 10.0-18.1) times as likely to be HIV-infected as women of reproductive age [5-6]. In 2018 in Guigang City, Guangxi China, HIV prevalence among FSWs was 11.8 %[29]. While prevalence amongst FSWs remained high at 6.2% (95% CI: 4.4-8.3) with a relatively low national HIV prevalence of 0.6% in Brazil [26]. However in 2020, there were 1.5 [1.0-2.0] million new HIV infections worldwide. Declines in new HIV infections among adults have slowed in recent years, from 3.0 [2.1-4.2] million to 1.5 [1.0-2.0] million new infections[26], and this calls for refurbished HIV prevention measures to reduce the incidence of HIV.

Sub-Saharan Africa remains most severely affected, with nearly 1 in every 25 adults (4.2%) living with HIV and accounting for nearly two-thirds(67%) of the people living with HIV worldwide [26]. In 2017, a study mapped female sex workers in Kenya; 3.0% of the adult female population in Nairobi constituted of female sex workers in 2017, the FSW prevalence was 4.8%[23]. The highest prevalence of FSWs was seen in Madagascar Diego-Suarez Provincial town at 12.0 % [26].The pooled HIV prevalence among Female Sex Workers in the west and central Africa was 34.9% (*n=*14,388/41,270), among their clients was 7.3%. HIV prevalence was high among a sample of FSWs in Rwanda; the overall prevalence was 51% [11-12].

In Uganda, approximately 1.3 million people aged 15 to 64 are living with HIV; HIV prevalence in the adult population was 5.5% (7.1 % and 3.8% in women and men, respectively) in 2020[21-25]. It has been shown that HIV rates are increasing among young females aged (15-24) at 2.9% compared to young males at 0.8% in Uganda[21-25]. HIV prevalence is almost three times higher among females than males aged 15 to 19 and 20 to 24 [21]. The HIV epidemic in Uganda continues to disproportionately affect women especially young women, according to the Ministry of Health, 2020 preliminary results, HIV prevalence in women of reproductive age is 7.1% compared to 3.8% among males of reproductive age[21].HIV prevalence is higher among women living in urban areas (9.8%) than those in rural areas (6.7%), attributed to increased commercial sex work [21]. Similarly, 570 young women aged 15-24 acquire HIV every week in Uganda [25], attributed to insufficient sexual education. A study conducted in 2014, revealed that 38.5% of young women and men aged 15-24 could correctly identify ways of preventing ttransmission of HIV and rejected major misconceptions about HIV transmission[12]. HIV prevalence among female sex workers in Kampala is 33% [18-21]. Common drivers of the HIV epidemic among female sex workers include Intimate partner violence, stigma, discrimination, and laws against sex work [19]. Intimate partner violence contributes to high HIV prevalence among sex workers because male perpetrators of violence are more likely to be HIV positive, when there is sexual violence there is a reduced capacity of the sex worker to negotiate condom use, sexual violence increases tearing and wounds during sex which increases the risk of acquiring HIV and STIs [26]. Although TDF-based PrEP can reduce the risk of HIV infection if taken 72 hours after exposure, there are some gaps in PrEP knowledge, and these include delivery models to maximize uptake and retention[22]. The data available is not conclusive.

Therefore, the main objective of this study was to determine factors influencing retention of FSWs at 1 month on PrEP in the three health facilities of MARPI, Kisenyi and Kawala. We hypothesized that client characteristics, social and drug-related factors might adversely affect retention of Female sex workers at 1 month. Understanding factors that influence retention of FSWs at 1 month on PrEP is important in generating evidence that might inform MOH and implementing partners in designing interventions for improved PrEP service delivery among the Female sex workers.

## 2. Materials and Methods

The study was conducted using data from the Most at Risk Populations Initiative (MARPI) clinic located within Mulago National Referral Hospital, Kisenyi and Kawala HCs in central Uganda. These sites offer PrEP to FSWs and other key populations: MARPI is a Centre’s for Disease Control and Prevention[9] funded programme that started in 2008 to specifically deal with health issues affecting key populations, including FSWs that are at a high risk of contracting the HIV virus. Kisenyi and Kawala HCs are public health facilities that offer KP-friendly health services.

We used a retrospective cohort study design because we had to follow up clients who had been initiated on PrEP one month ago. We conducted in-depth interviews with thirty FSWs among those adhering well to PrEP, and always coming for refills, among those who ever missed coming for PrEP refill but later came back for the refill and among those who stopped PrEP completely and stopped getting PrEP refills.

To ensure that each female sex worker using PrEP had equal opportunities for inclusion to participate in the study, a systematic sampling procedure was used to get the FSWs that would be interviewed. Only the first unit was selected randomly, otherwise, every 2^nd^ FSW who came for a PrEP refill at the clinic was interviewed. Two research assistants were trained to collect data among illiterate and semi-illiterate FSWs using researcher-administered semi-structured questionnaires in Luganda.

Before the interviews, the semi-structured questionnaire was translated from English to Luganda (One translated from English to Luganda and the other back translated from Luganda to English), by two independent local translators, two for each language. Thereafter, the two translators for each language jointly harmonized the final questionnaires to facilitate comprehension among the illiterate and semi-illiterate FSWs. Prior to data collection, we obtained written consent from all the study participants. On daily basis, the research assistants and principal investigator reviewed the questionnaires for accuracy and completeness. Data was collected from Monday to Friday because PrEP clinics are run only on weekdays.. Quantitative data were double entered into Epi-Data version 3.1 and exported to STATA version 14.0 for univariate, bivariate and multivariate analysis at 5% significance level.

Independent variables were client characteristics (age, education level, religion, work place, marital status and use of lubricants), social factors (attitude of health workers, stigma, disclosure, lack of family support and knowledge of PrEP among FSWs), Drug-related factors (pill color and size, perceived side effects, pill packaging, cost of drugs and availability of PrEP drugs).Retention was measured through establishing the number of days off PrEP drugs

In Univariate analysis, we computed the frequencies and percentages for categorical variables. In Bivariate analysis, the Poisson regression model was used to determine the association between independent variables and the outcome variable.The results were reported into unadjusted Incidence Rate Ratios (UIRR).

For all significant results at bivariate analysis, were analysed using the Poisson regression model to determine factors that influence retention of FSWs on PrEP at 1 month, and results were reported as Adjusted Incidence Rate Ratios (AIRR). In all analyses at Univariable and Multivariable poisson regression, all Incidence Rate Ratios were reported with their corresponding 95% Confidence Interval.

This study was approved by the Institutional Review Board of Clarke International University (Formerly International Health Sciences University), Institute of Public Health and Management, Kampala Uganda with a reference number Clarke-2021-211.

## 3. Results

### Client characteristics

In total 350 female sex workers were interviewed and the median age of the FSWs was 25 years. 40.9% of the FSWs had attended primary level, 43.7% secondary level, 3.4% tertiary level education and 12% had no education. (table 1).

**Table 1:** shows the characteristics of the study participants;

| Variable | Category | N (%) |
| --- | --- | --- |
| Age(yrs) | 18-19 | 45(13) |
|  | 20-29 | 207(59) |
|  | 30-39 | 88(25) |
|  | 40+ | 10(3) |
| Marital status | Divorced | 115(32.86) |
|  | Married | 31(8.86) |
|  | Single | 200(57.14) |
|  | Widowed | 4(1.14) |
| Religion | SDA | 2(0.6) |
|  | Catholic | 134(38.3) |
|  | Muslim | 83(23.7) |
|  | Pentecostal | 61(17.4) |
|  | Protestant | 70(20.0) |
|  | Student | 5(1.4) |
| Education level | None | 42(12.0) |
|  | Primary | 143(40.9) |
|  | Secondary | 153(43.7) |
|  | Tertiary | 12(3.4) |

The mean duration of sex work was 3 years. On average, the FSWs had 14 sex partners in the past week. 59.4% reported lubricant use on the last sexual encounter compared to 40.6% who did not use lubricants(Table 2)

**Table 2:** shows the social factors that determine retention of FSWs on PrEP at I month

| Variable | Category | Frequency | Percentage |
| --- | --- | --- | --- |
| Do you walk to the health facility to access your PrEP refills | No | 152 | 43.4 |
|  | Yes | 198 | 56.6 |
| Kilometres walked | Less than 5 km | 128 | 64.7 |
|  | 5-10 km | 65 | 32.8 |
|  | Above 10 km | 5 | 2.5 |
| Use boda boda to access Health Facility | Yes | 168 | 48 |
|  | No | 182 | 52 |
| Amount spent on transport to access PrEP | 3000 - 5000 | 75 | 44.4 |
|  | 5500 - 8000 | 45 | 26.8 |
|  | 8500 - 12000 | 30 | 17.9 |
|  | 12500 - 15000 | 11 | 6.6 |
|  | Over 15000 | 7 | 4.2 |
| sex being main source of income | No | 23 | 6.6 |
|  | Yes | 327 | 93.4 |
| Place of sex work | Brothel | 141 | 40.3 |
|  | Home | 79 | 22.6 |
|  | Street | 130 | 37.1 |
| Disclosure to sexual partner that you are on PrEP | No | 233 | 66.6 |
|  | Yes | 117 | 33.4 |
| Disclosure to a family that is FSW on PrEP | No | 231 | 66 |
|  | Yes | 119 | 34 |
| Have you been stigmatised | No | 184 | 52.6 |
|  | Yes | 166 | 47.4 |
| condom use with a paying customer | No | 59 | 16.9 |
|  | Yes | 291 | 83.1 |
| condom use with a non-paying customer | No | 157 | 44.9 |
|  | Yes | 193 | 55.1 |
| use of lubricant on last sexual encounter | No | 142 | 40.6 |
|  | Yes | 208 | 59.4 |
| Would lubricants and condoms make you come for PrEP refills (on scale 1-10) | 1 | 16 | 4.57 |
|  | 2 | 25 | 11.71 |
|  | 3 | 16 | 4.57 |
|  | 4 | 22 | 6.29 |
|  | 5 | 41 | 11.71 |
|  | 6 | 32 | 9.14 |
|  | 7 | 39 | 11.14 |
|  | 8 | 50 | 14.29 |
|  | 3 | 38 | 10.86 |
|  | 10 | 71 | 10.86 |
| Alcohol consumption | No | 114 | 32.6 |
|  | Yes | 236 | 67.4 |
| Alcohol consumption on nights/days of selling sex | No | 114 | 32.6 |
|  | Yes | 236 | 67.4 |
| Type of alcohol used | beer 500mls | 93 | 31.2 |
|  | beer 350mls | 105 | 35.2 |
|  | Waragi bottles | 29 | 9.7 |
|  | Waragi sachets | 66 | 22.2 |
|  | local brew | 5 | 1.9 |
| use of drugs | No | 264 | 75.4 |
|  | Yes | 86 | 24.6 |
| Drug use on nights/days of selling sex | Yes | 86 | 100 |
| Forgetting PrEP clinic day due to drugs/alcohol | No | 224 | 74.7 |
|  | Yes | 76 | 25.3 |
| Counselled about PrEP before initiation | No | 17 | 4.6 |
|  | Yes | 333 | 95.4 |

**Table 3:**
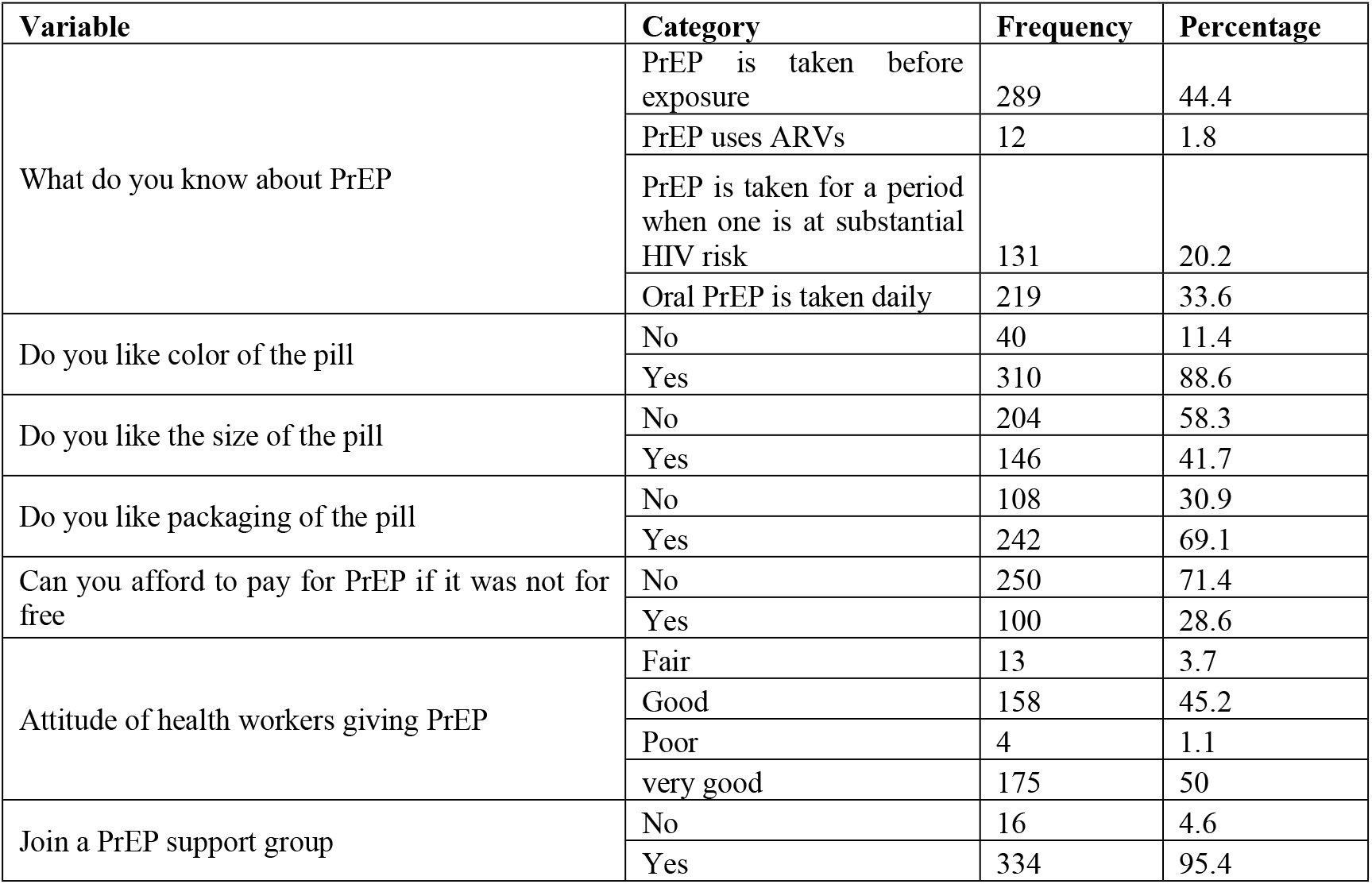
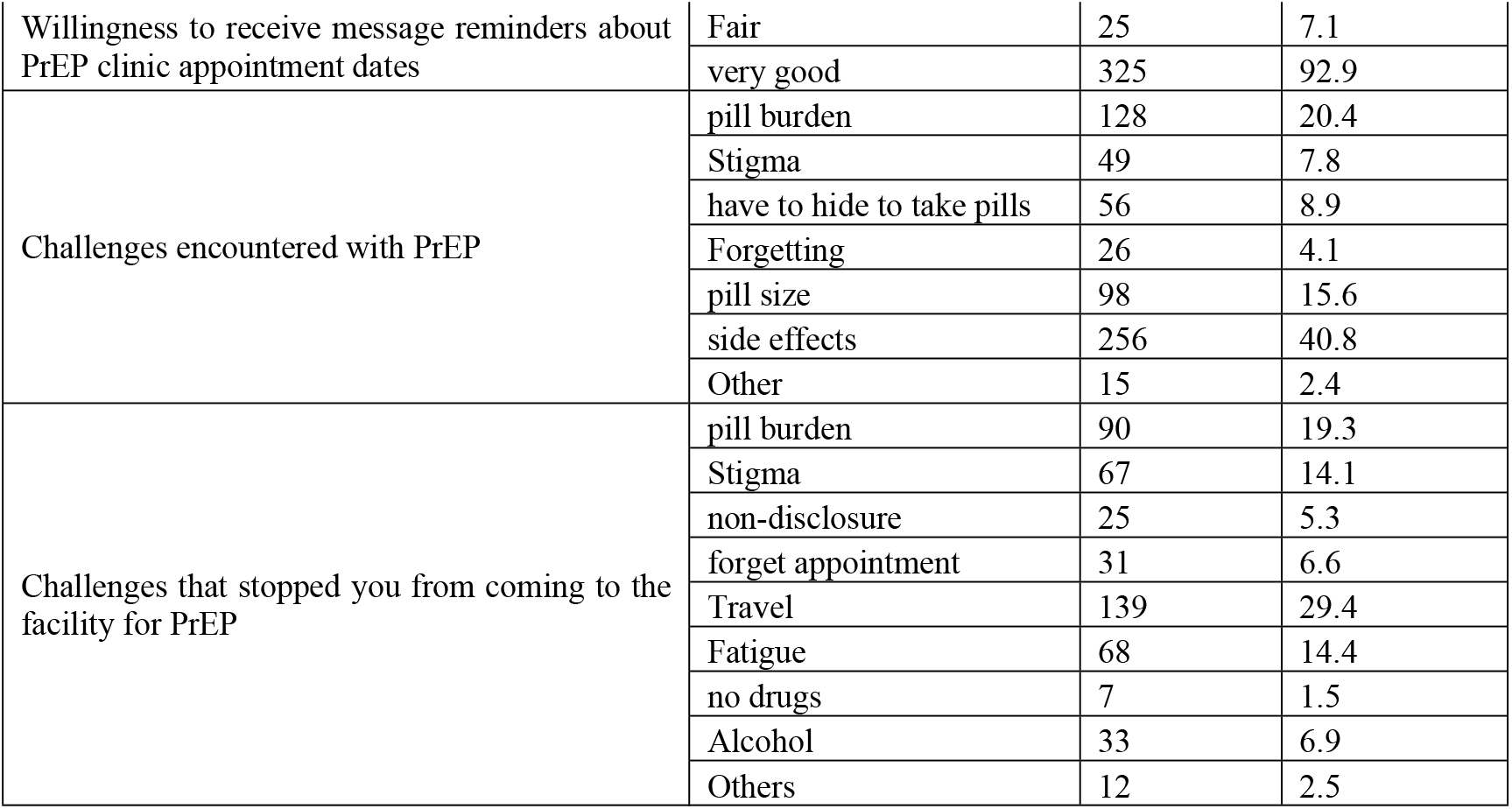
shows the drug related factors that determine retention of FSWs on PrEP at I month

### Determinants of one month PrEP Retention of FSWs

In bivariate analysis, Retention of FSWs on PrEP was 38% times higher among the protestant sex workers ((p=0.041, [1.013, 1.88]) and 73% times higher among the Pentecostal sex workers (p=0.051, [0.999, 1.855]).Place of work (p=0.02, [1.01, 1.11]), retention was 6% times higher among sex workers that work from the street as compared to those that work in brothels.

Retention among the sex workers that use lubricant was 4% times lower than that among those that did not use lubricant (p=0.006, [1.02,1.10]), and for those who use drug it was equally 4% times lower than that among those that did not use drugs (p=0.010, [0.897,0.986]).

Retention among FSWs who received PrEP counselling was 25% times higher than that among those that did not receive counselling (p<0.001, [1.12, 1.40]). Colour of PrEP tablet (p=0.007, [1.03, 1.17]) not liking the colour of PrEP was negatively associated with high PrEP retention, whereby FSWs who did not like the colour of PrEP was 90.6% less likely to be retained to PrEP; FSWs who did not like the PrEP packaging (p<0.001, [1.07, 1.17]) whereby not liking the PrEP packaging was negatively associated with retention to PrEP. FSWs who did not like the PrEP packaging were 88% less likely to be retained to PrEP. PrEP Pill size (p=0.002, [1.02, 1.11]), retention among FSWs that liked the size of the PrEP pill was 7% times higher than that those that did not like the pill size. Attitude of health workers (p=0.002, [1.07, 1.35]), attitude of health workers was positively associated with retention of FSWs on PrEP, retention was 20% higher among the sex workers that had perceived that the health care attitude to be good compared those that perceived to be fair(table 4).

**Table 4:**
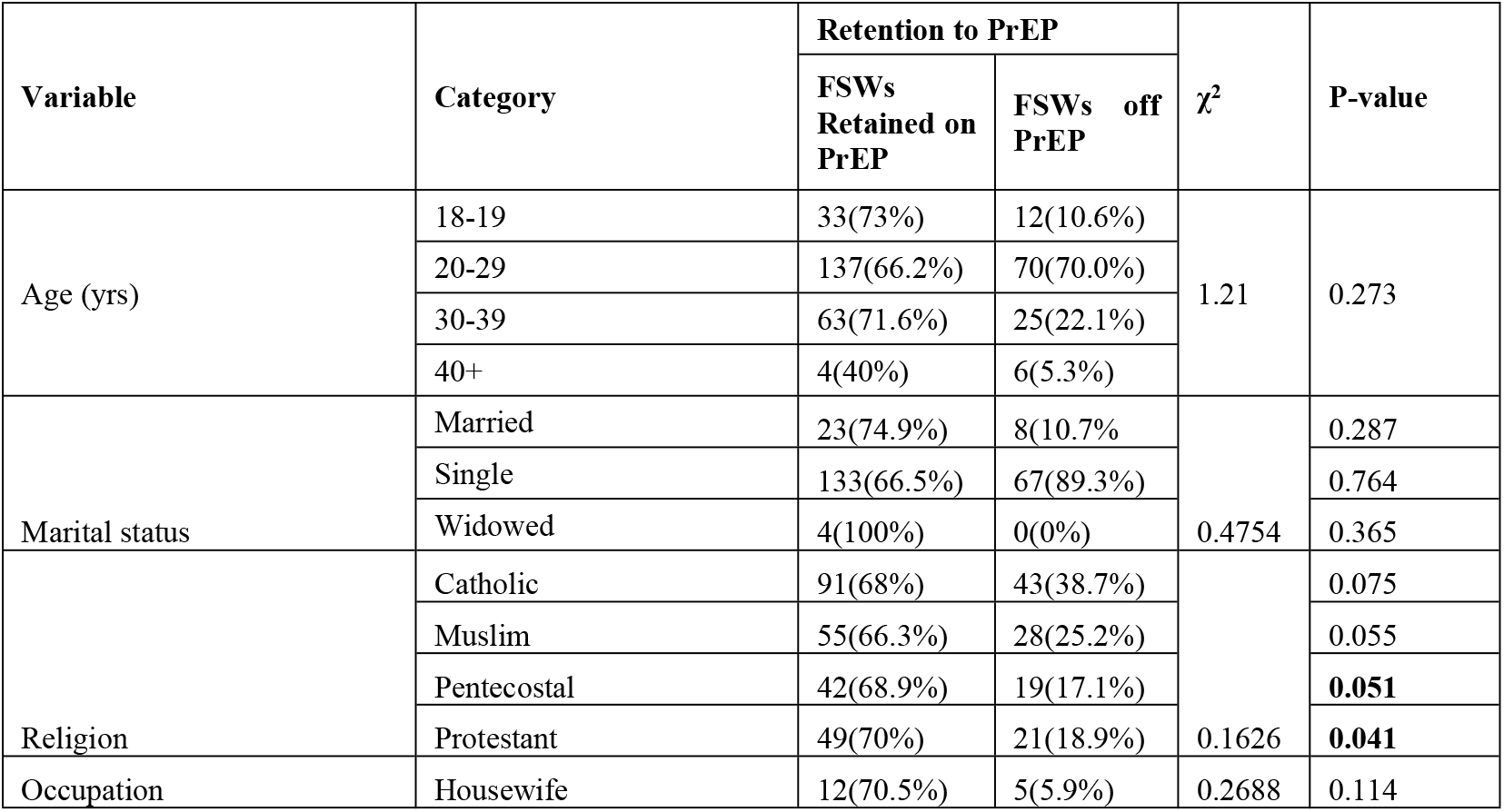
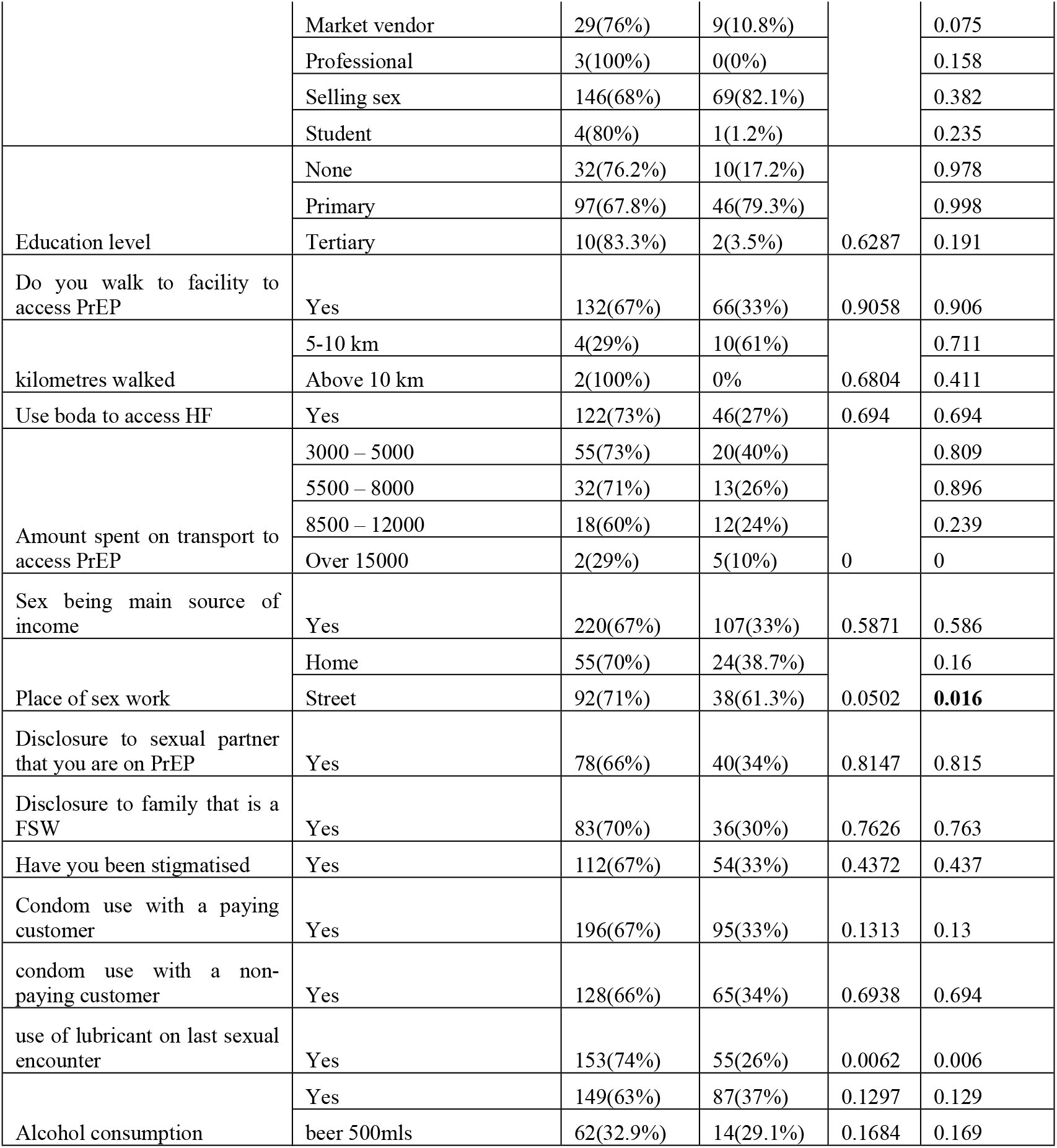
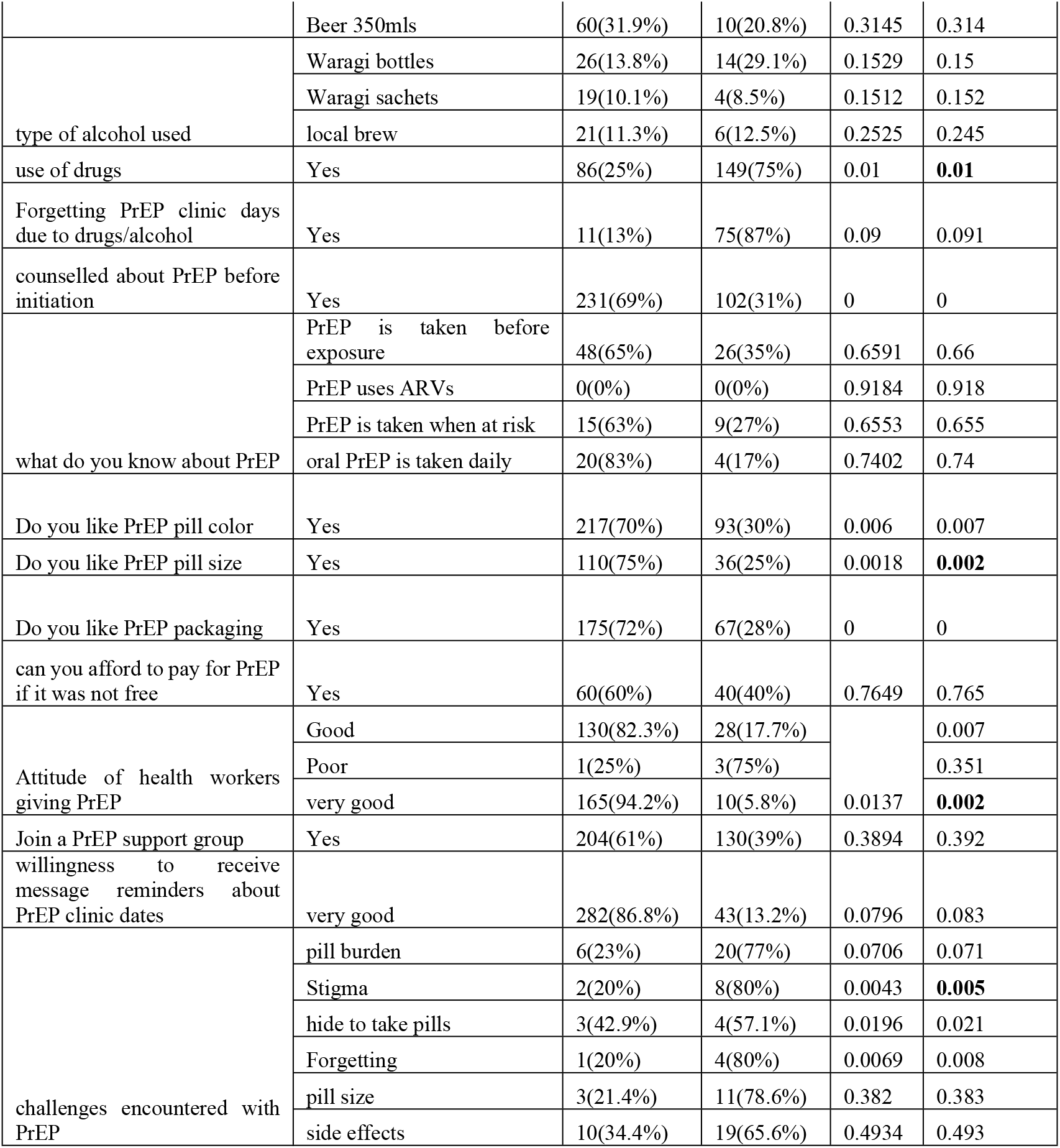
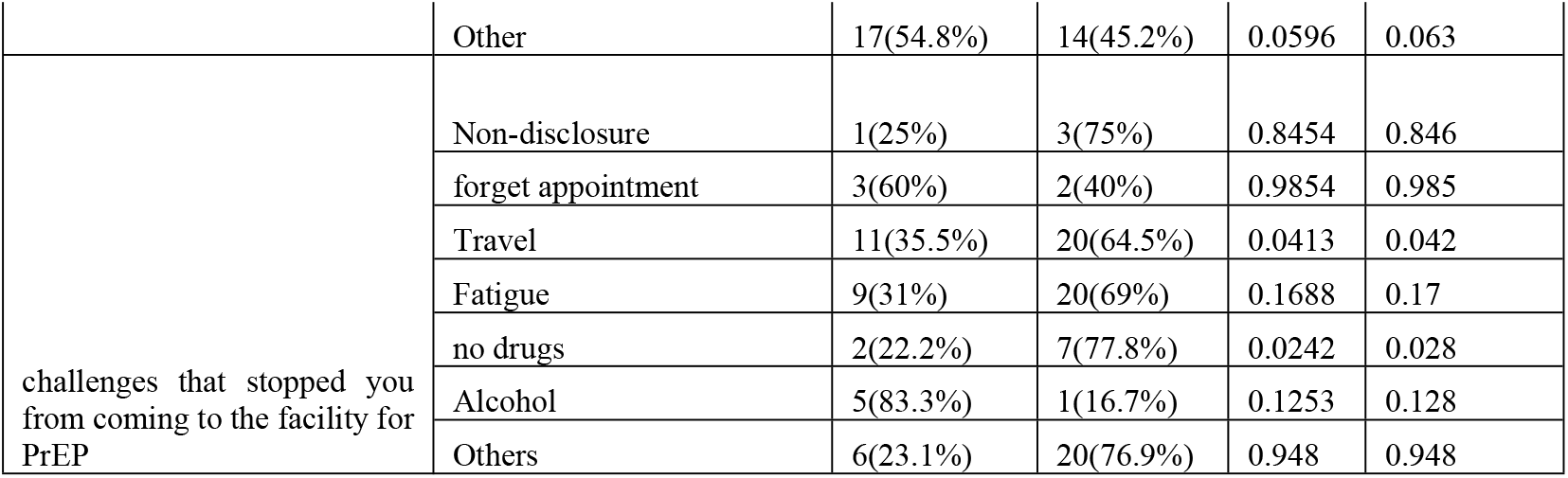
Bivariate Analysis of factors associated with retention of FSWs to PrEP at 1 month.

The major challenges the FSWs encountered in 1 month of taking PrEP were: stigma (p=0.005, [IRR 8% lower]), hide to take pills (p=0.02, [IRR 7% lower]), forgetting (p=0.008, [IRR 10% lower]). Retention to PrEP is 8% lower among sex workers for every passing days of having sex in the past week (p<0.005, [1.02, 1.04]).For FSWs that had stopped coming for PrEP refills at the facility the challenges they had were; no drugs(p=0.028, [IRR 15% lower]) and travel (p=0.04, [IRR 4% lower]).

In Multivariable poisson regression (adjusted for all significant variables at univariate analysis), use of lubricants, health workers’ attitude perceived as good, stigma and PrEP/STI drug stock outs were significantly associated with retention of FSWs on PrEP at 1 month. However, pill color and size, forgetfulness, hiding to take drugs and use of drugs were not associated with retention of FSWs to PrEP (Table 5).

**Table 5:** Multivariate Analysis of factors associated with retention to PrEP among FSW at 1 month.

| Variable | Category | Retention to PrEP |  | IRR | 95%CI | P-value |
| --- | --- | --- | --- | --- | --- | --- |
|  |  | FSWs Retained on PrEP | FSWs off PrEP |  |  |  |
| Use of lubricants | Yes | 153(74%) | 55(26%) | 1.07 | [1.01, 1.14] | **0.02** |
| Use of drugs | Yes | 86(25%) | 149(75%) | 0.97 | [0.91, 1.03] | 0.39 |
| PrEP pill color | Yes | 217(70%) | 93(30%) | 1.05 | [0.95, 1.17] | 0.27 |
| PrEP pill size | Yes | 110(75%) | 36(25%) | 1.04 | [0.99, 1.11] | 0.11 |
| Health workers' attitude | Good | 130(82.3%) | 28(17.7%) | 1.23 | [1.05, 1.44] | **0.012** |
|  | Poor | 1(25%) | 3(75%) | 1.31 | [0.93, 1.87] | 0.12 |
|  | very good | 165(94.2%) | 10(5.8%) | 1.24 | [1.06, 1.46] | 0.006 |
| Challenges encountered with PrEP | Stigma | 2(20%) | 8(80%) | 0.9 | [0.814, 0.996] | **0.04** |
|  | Hide to take pills | 3(42.9%) | 4(57.1%) | 1 | [0.92, 1.09] | 0.98 |
|  | Forgetting | 1(20%) | 4(80%) | 0.93 | [0.83, 1.04] | 0.208 |
| Challenges that stopped you from coming to the facility for PrEP | Travel | 11(35.5%) | 20(64.5%) | 0.98 | [0.93, 1.04] | 0.57 |
|  | No drugs | 2(22.2%) | 7(77.8%) | 0.76 | [0.67, 0.76] | **0.003** |

## 4. Discussion

Whereas retention of FSWs to PrEP at 1 month in this study was sub-optimal (68%), it was attributed to lack of enticing commodities such as STI drugs and lubricants at the health facilities, PrEP drug stock outs, pill burden and drug side effects. This findings are consistent with other literature about FSWs in Kampala that were studied in an earlier study [18].

Additionally, it is important to note that from the IDIs, FSWs indicated that they had a lot of misconceptions and myths about PrEP due to inadequate counselling prior to initiation by the health workers that led to many FSWs dropping off the program. Relatedly from the qualitative interviews, PrEP packaging led to stigma among the FSWs.This is probably because FSWs were stigmatised by their colleagues that they were HIV positive and taking ART due to similar packaging of PrEP drugs.

This study found out that there is a significant relationship between use of lubricants and retention among FSWs (adjusted 0.02 [1.01, 1.14] IRR 1.07).FSWs who use lubricants were more retained on PrEP.Relatedly, FSWs preferred to be given PrEP from where they work (hotspots, brothels and the DICs), however they were not reached by the outreach team, which had not yet been boosted at that time. It is also important to note that participants were not reimbursed their money which they used for transport and other expenditures on a clinic day, which is probably why many FSWs in the qualitative interviews mentioned that they preferred getting PrEP from the community/outreach, and why those who dropped out of care explained in the in-depth interviews that they found it challenging to come back to the clinic because they find it hard to travel for PrEP refills, remembering that most of these FSWs have joined sex work because of a poor socio-economic status(such as being widowed,illiterate,orphaned at an early age) and from this study findings, FSWs earn a monthly mean income of 203,200 shillings a month.The study findings show that retention was higher among FSWs who had a perception that the health workers’ attitude was good (adjusted 0.012 [1.05, 1.44] IRR 1.23 and among those who were stigma free) (adjusted 0.04 [0.814, 0.996] IRR 0.90) with adherence of 82%.The iPrEx modelling study which reflected data from MSM PrEP users showed that the estimated PrEP efficacy was 76% for MSM who took 2 doses per week, 96% for 4 doses per week, and 99% for 7 doses per week. However, the pharmacokinetics differ for other routes of HIV transmission (anal versus vaginal)[1-2]. These findings indicate that the high level of PrEP adherence and retention achieved in the setting of a PrEP implementation site can be associated with a high level of protection against HIV acquisition by the HIV-uninfected female sex workers in comparison with the iPrEX study findings (The mean number of days for taking PrEP increased by 2.7% for every passing day of having sex in the past week).

This study found out that, retention to PrEP was lower among FSWs who did not receive counselling before initiation(p<0.001, [1.12, 1.40]). In this case, counsellors need to emphasize the need for the FSWs what is PrEP, the benefits, side effects, duration of taking PrEP, adherence to PrEP and the need to be retained in care.

This finding was in agreement with a study done by mathematical modelling to compare the impact of increasing condom use or HIV pre-exposure prophylaxis (PrEP) use among female sex workers (Mukandavire, 2016). The study equally found out that, PrEP retention among FSWs who use drugs was lower as compared to those who do not use drugs(p=0.010, [0.897,0.986]) due to being drunk on the clinic appointment days or having forgotten the clinic appointment days.

Retention of FSWs on PrEP at 1 month in selected health facilities Kampala was not significantly associated with disclosure and family support.

No drugs at the facility affected PrEP retention among FSWs(adjusted 0.003 [0.67, 0.922] IRR 0.76).This is slightly lower than PrEP retention levels from another study that offered PrEP to FSWs in South Africa where PrEP retention ranged from 75% to 85% [15]. The retention was probably higher in this study in South Africa whereby PrEP was offered to FSWs in a demonstration study setting while in this study in Kampala PrEP was offered at public health facilities whereby there was no transport reimbursement given to PrEP clients.

‘PrEP packaging’ and ‘color of PrEP’ were negatively associated with retention to PrEP.FSWs who did not like the PrEP packaging were 88% less likely to be retained to PrEP as recommended compared to those FSWs who reported that they liked the PrEP packaging (p<0.001, [1.07, 1.17]). FSWs who did not like the color of PrEP was 90.6% less likely to be retained to PrEP (p<0.007, [1.03, 1.17]). Retention among FSWs that liked the size of the PrEP pill was 7% times higher than that those that did not like the pill size (p<0.002, [1.02, 1.11]). This was supported by the qualitative findings whereby some people mentioned that the pills were just like ARVs in terms of the size, color and packaging and they did not like PrEP similarity to ART. Many mentioned that they had to hide their pill bottles, and thus just moved with a few pills in their bras or others re-packaged their pills into smaller disposable polythene bags.

Relatedly, a study among women in South Africa showed that pill characteristics of PrEP affect retention to PrEP whereby liking the pill color (OR: 2.93; 95% CI: 1.18-7.27) was positively associated with good retention [12-13]. In contrast, a study done in Botswana found that pill count adherence was significantly associated with adverse events (nausea, dizziness, vomiting) (RR 0.98 95 % CI 0.98-1.00; p = 0.03) and side effect concerns (RR 0.98 95 % CI 0.96-0.99; p = 0.01) [19]. This was not found significant in the study among FSWs in Kampala. The patterns of visits to the clinic for PrEP, discovered from the people, who according to the client register were described as ‘dropped out of care’, yet on interviewing them, they mentioned that they ‘later resumed taking PrEP’ after some time, combined with the reported adherence level suggest intermittent use of PrEP, whereby FSWs take a break from PrEP for different reasons such as ‘when they feel that they are tired of the tablets’, or when ‘they have fewer customers’ and when their ‘customers are using condoms’, or when they take a break from sex work due to family problems such as the illness of a relative, burials, travel et cetera while maintaining an overall engagement with HIV prevention services.

This study found that 92.8% of the participants would like to receive a message to remind them about their clinic appointments, if this is feasible it is a good and cheap way to improve appointment keeping and retention to PrEP because even during the qualitative interviews many participants wished for a form of communication from the health workers to reassure them when they experienced side-effects, and to keep reminding and encouraging them to take PrEP. Similarly, a randomized controlled trial of text message reminders found that mobile phone technologies improve adherence to antiretroviral treatment in a resource-limited setting (Pop-Eleches et al., 2011). They found that participants in groups receiving weekly reminders were significantly less likely to experience treatment interruptions exceeding 48 hours during the 48-week follow-up period than participants in the control group (81 vs. 90%, *P* = 0.03)(Pop-Eleches et al., 2011).The key to note is that the cost of PrEP drugs was not significantly associated with retention of FSWs on PrEP.

## 5. Study Limitations

This study was subject to some limitations. While we interviewed some clients, who were no longer coming for PrEP refills, several others were inaccessible having travelled abroad or moved to a district in Northern, Western or Eastern Uganda and thus were not included in our sample. However, some were asked if they could at least disclose the reason for their failure to return to the clinic to get PrEP refills. Some of the phone numbers of those FSWs who were not coming for PrEP refills were off for long periods, and other phone numbers were no longer in service due to the mobile nature of FSWs who had travelled to upcountry places for business. The findings therefore do not represent the experiences of these clients who were no longer coming for PrEP refills, and their reasons for dropping out of care. And lastly, some women who were recorded as having missed PrEP scheduled visits in the clinic register, when contacted, said that they were not sex workers and that they have never been sex workers.

## 6. Conclusion

Our study shows that FSWs get retained in care on PrEP in the Public health facilities of Kampala when PrEP is enhanced with enticing commodities such as lubricants and STI drugs, are counselled before PrEP initiation thus stigma-free and health workers’ attitude is perceived good.

Interventions should thus focus on training health workers’ on Key population friendly services, repetitive PrEP education to clear the myths and misconceptions and provision of enticing commodities to enhance PrEP.

## Data Availability

The datasets generated and/or analysed during the current study are not publicly available due to the dataset containing sensitive personal data. Aggregated data and mixing matrices data are available from the corresponding author on reasonable request

## Ethics statement

Our study received ethical review and approval from Clarke International University Research Ethics Committee (CIU-REC with a reference number Clarke-2021-211). We obtained administrative approval from the Public Health Directorate of Kampala Capital City Authority.

## Acknowledgments

We are grateful to the health facility heads of the respective study sites and the Public Health department of Kampala Capital City Authority for their unwavering support.

## Author’s contribution

James Wanyama and John Bosco Alege designed the study. All authors (except Christine Atuhairwe) participated in the development and validation of the study questionnaires. James Wanyama and John Bosco Alege drafted the article and supervised the data collection and analysis. James Wanyama and John Bosco Alege participated in data collection. James Wanyama,John Bosco Alege and Christine Atuhaire participated in the analysis. All authors interpreted the results and critically revised the manuscript for scientifc content. All authors approved the final version of the article

## References

[1] Andersen, R. and Newman, J.F. (2005) “Societal and Individual Determinants of Medical Care Utilization in the United States,” Milbank Quarterly, 83(4), p. Online-only-Online-only. doi:10.1111/j.1468-0009.2005.00428.x.

[2] Anderson, P.L. et al. (2012) “Emtricitabine-Tenofovir Concentrations and Pre-Exposure Prophylaxis Efficacy in Men Who Have Sex with Men,” Science Translational Medicine, 4(151). doi:10.1126/scitranslmed.3004006.

[3] Auerbach, J.D. et al. (2015) “Knowledge, Attitudes, and Likelihood of Pre-Exposure Prophylaxis (PrEP) Use Among US Women at Risk of Acquiring HIV,” AIDS Patient Care and STDs, 29(2), pp. 102–110. doi:10.1089/apc.2014.0142.

[4] Baeten, J.M. et al. (2012) “Antiretroviral Prophylaxis for HIV Prevention in Heterosexual Men and Women,” New England Journal of Medicine, 367(5), pp. 399–410. doi:10.1056/NEJMoa1108524.

[5] Baral, S. et al. (2012) “Burden of HIV among female sex workers in low-income and middle-income countries: a systematic review and meta-analysis,” The Lancet Infectious Diseases, 12(7), pp. 538–549. doi:10.1016/S1473-3099(12)70066-X.

[6] Baral, S.D. et al. (2013) “Worldwide burden of HIV in transgender women: a systematic review and meta-analysis,” The Lancet Infectious Diseases, 13(3), pp. 214–222. doi:10.1016/S1473-3099(12)70315-8.

[7] Bekker, L.-G. et al. (2015) “Combination HIV prevention for female sex workers: what is the evidence?,” The Lancet, 385(9962), pp. 72–87. doi:10.1016/S0140-6736(14)60974-0.

[8] Bukenya, J. et al. (2013) “Condom use among female sex workers in Uganda,” AIDS Care, 25(6), pp. 767–774. doi:10.1080/09540121.2012.748863.

[9] CDC (2018) Pre-exposure prophylaxis for the prevention of HIV infection in the United States -2014: a clinical practice guideline.

[10] Chakrapani, V. et al. (2015) “Acceptability of HIV Pre-Exposure Prophylaxis (PrEP) and Implementation Challenges Among Men Who Have Sex with Men in India: A Qualitative Investigation,” AIDS Patient Care and STDs, 29(10), pp. 569–577. doi:10.1089/apc.2015.0143.

[11] Chersich, M.F. et al. (2013) “Priority interventions to reduce HIV transmission in sex work settings in sub-Saharan Africa and delivery of these services,” Journal of the International AIDS Society, 16(1), p. 17980. doi:10.7448/IAS.16.1.17980.

[12] Corneli, A.L. et al. (2014) “FEM-PrEP,” JAIDS Journal of Acquired Immune Deficiency Syndromes, 66(3), pp. 324–331. doi:10.1097/QAI.0000000000000158.

[13] Cowan, F.M. and Delany-Moretlwe, S. (2016) “Promise and pitfalls of pre-exposure prophylaxis for female sex workers,” Current Opinion in HIV and AIDS, 11(1), pp. 27–34. doi:10.1097/COH.0000000000000215.

[14] Dayer, L. et al. (2013) “Smartphone medication adherence apps: Potential benefits to patients and providers,” Journal of the American Pharmacists Association, 53(2), pp. 172–181. doi:10.1331/JAPhA.2013.12202.

[15] Eakle, R. et al. (2018) “Exploring acceptability of oral PrEP prior to implementation among female sex workers in South Africa.” doi:10.1002/jia2.25081/full.

[16] Fonner, V.A. et al. (2016) “Effectiveness and safety of oral HIV preexposure prophylaxis for all populations,” AIDS, 30(12), pp. 1973–1983. doi:10.1097/QAD.0000000000001145.

[17] Glynn, J.R. et al. (2001) “Why do young women have a much higher prevalence of HIV than young men? A study in Kisumu, Kenya and Ndola, Zambia,” AIDS, 15, pp. S51–S60. doi:10.1097/00002030-200108004-00006.

[18] Hladik, W. et al. (2017) “Burden and characteristics of HIV infection among female sex workers in Kampala, Uganda – a respondent-driven sampling survey,” BMC Public Health, 17(1), p. 565. doi:10.1186/s12889-017-4428-z.

[19] Kebaabetswe, P.M. et al. (2015) “Factors Associated with Adherence and Concordance Between Measurement Strategies in an HIV Daily Oral Tenofovir/Emtricitibine as Pre-exposure Prophylaxis (Prep) Clinical Trial, Botswana, 2007–2010,” AIDS and Behavior, 19(5), pp. 758–769. doi:10.1007/s10461-014-0891-z.

[20] Khawcharoenporn, T., Kendrick, S. and Smith, K. (2012) “HIV Risk Perception and Preexposure Prophylaxis Interest Among a Heterosexual Population Visiting a Sexually Transmitted Infection Clinic,” AIDS Patient Care and STDs, 26(4), pp. 222–233. doi:10.1089/apc.2011.0202.

[21] MINISTRY OF HEALTH, Uganda. (2019) Population-based HIV Impact Assessment (UPHIA) 2020-2021: Final Report. Kampala.

[22] Muwonge TR, Nsubuga R, Brown C, et al. Knowledge and barriers of PrEP delivery among diverse groups of potential PrEP users in Central Uganda. PLoS One. 2020;15(10):e0241399. Published 2020 Oct 28. doi:10.1371/journal.pone.0241399

[23] Tago A, Mckinnon LR, Wanjiru T, Muriuki F, Munyao J, Gakii G, Akolo M, Kariri A, Reed N, Shaw SY, Gelmon LJ, Kimani J. Declines in HIV prevalence in female sex workers accessing an HIV treatment and prevention programme in Nairobi, Kenya over a 10-year period. AIDS. 2021 Feb 2;35(2):317–324. doi:

[24] Tangmunkongvorakul, A. et al. (2013) “Facilitators and barriers to medication adherence in an HIV prevention study among men who have sex with men in the iPrEx study in Chiang Mai, Thailand,” AIDS Care, 25(8), pp. 961–967. doi:10.1080/09540121.2012.748871.

[25] UGANDA AIDS COMMISSION (2015) National HIV and AIDS Strategic Plan 2015/2016-2019/2020, An AIDS free Uganda, My responsibility! Kampala.

[26] UNAIDS (2021) Global HIV & AIDS statistics — Fact sheet. Available at: https://www.unaids.org/en/resources/fact-sheet (Accessed: February 7, 2022).

[27] UNFPA (2006) 17 Fact Sheets with concise information on gender-related aspects of HIV/AIDS. Kampala.

[28] Vandepitte, J. et al. (2006) “Estimates of the number of female sex workers in different regions of the world.,” Sexually transmitted infections, 82 Suppl 3, pp. iii18–25. doi:10.1136/sti.2006.020081.

[29] Zhang, L. et al. (2015) “A systematic review and meta-analysis of the prevalence, trends, and geographical distribution of HIV among Chinese female sex workers (2000–2011): implications for preventing sexually transmitted HIV,” International Journal of Infectious Diseases, 39, pp. 76–86. doi:10.1016/j.ijid.2015.08.014.

